# Lymphatic Filariasis Transmission at Spot-Check Sites in Six Endemic Districts of Nepal After Two IDA Mass Drug Administration Rounds

**DOI:** 10.64898/2026.04.22.26351459

**Authors:** Ram Kumar Mahato, Gokarna Dahal, Shashi Kandel, Anish Chaudhary, Satya Raj Paudel, Rabindra Khaniya, Prakash Shakya, Bhim Prakash Devkota, Bhim Prasad Sapkota, Krishna Prasad Paudel, Bijay Bajracharya, Deena Shrestha, Chandra Bhal Jha, Roshan Neupane, Keshar Bahadur Dhakal, Kenza Bennani

**Affiliations:** Epidemiology and Disease Control Division, Teku, Kathmandu, Nepal; National Public Health Laboratory, Teku, Kathmandu; FAIRMED Foundation, Nepal; Department of Health Services, Teku, Nepal; Ministry of Health and Population, Ramshahpath, Kathmandu, Nepal; Center For Health and Disease Studies-Nepal; World Health Organization, Nepal

**Keywords:** Lymphatic Filariasis, Epidemiological Monitoring Survey, Mass Drug Administration

## Abstract

**Background:** Nepal has set a goal to eliminate lymphatic filariasis (LF) by 2030. As of 2024, Nepal has stopped the mass drug administration (MDA) in 56 of the 64 endemic districts and completed two rounds of MDA in six districts with persistent LF (≥2% antigen prevalence) using the three-drug regimen of Ivermectin, Diethylcarbamazine, and Albendazole (IDA), exceeding 65% coverage. We subsequently conducted an Epidemiological Monitoring Survey (EMS) to assess the impact of the MDA in reduction of LF infection prevalence below the transmission threshold and examine the factors associated with it.

**Methods:** We conducted a cross-sectional EMS nine months after MDA in 12 evaluation units (EUs) across six districts, with two sites per EU. We recruited a total of 7,343 individuals aged ≥20 years, sampled using multi-stage sampling, ensuring at least 300 blood samples collected per site. We collected data on demographics and MDA participation. We performed the LF antigen testing for all participants, followed by night blood microfilariae testing in antigen-positive individuals. Statistical analyses included non-parametric tests, Chi-square and Fisher’s Exact tests, and multivariable logistic regression to assess outcomes after adjusting for potential confounders.

**Results:** Nine of 12 evaluation units (EUs) recorded <1% microfilaremia, meeting the WHO threshold for passing EMS, while three EUs failed with ≥1% prevalence in at least one site. Antigen and MF prevalence were 4.47% and 0.34%, respectively (ratio 13:1). Both Antigen and MF prevalences were significantly associated with female sex (AOR= 0.564, 95% CI: 0.441–0.721 and AOR = 0.326, 95% CI: 0.129–0.826 respectively) and participation in the most recent MDA round (AOR = 0.477; 95% CI: 0.385–0.591 and AOR = 0.089; 95% CI: 0.017–0.464 respectively). MDA uptake was influenced by age (<40 years, AOR = 0.72; 95% CI: 0.653– 0.793), sex (female, AOR = 1.438; 95% CI: 1.29–1.603), cross-border residence (AOR = 0.616; 95% CI: 0.558–0.681), and occupation (agriculture and housewife, AOR = 1.144; 95% CI: 1.008–1.298). MF prevalence was also associated with younger age (<40 years, AOR = 0.211; 95% CI: 0.071–0.626).

**Conclusion:** The survey indicates progress toward LF elimination, with nine of twelve EUs achieving WHO’s <1% microfilaremia threshold after two rounds of IDA-MDA. However, transmission persists in three sites, likely linked to poor MDA participation among specific subgroups— particularly males, younger adults, and cross-border populations. Strengthening MDA coverage and compliance across all demographic and occupational groups, with special focus on border areas, is essential to achieve LF elimination in Nepal.

## Introduction

Lymphatic Filariasis (LF) is one of the neglected tropical diseases (NTD), caused by *Wuchereria bancrofti* and transmitted by Culex mosquitoes(1). This disease remains a public health challenge in many tropical and subtropical countries around the world(2). Globally, as of 2024, LF affects over 120 million people in 72 countries and is the second leading infectious cause of long-term infectious disability and imposes a significant socio-economic burden(3). Recognizing its public health importance, the World Health Organization (WHO) launched the Global Programme to Eliminate Lymphatic Filariasis (GPELF) in 2000 with the goal to eliminate LF as a public health problem initially by 2020 and later the goal was extended to 2030(4,5). The countries initially started Mass Drug Administration (MDA) using the Diethylcarbamazine and Albendazole to treat community people at risk and later adopted 3 drug regimens using the Ivermectin, Diethylcarbamazine, and Albendazole (IDA) to accelerate the elimination status as per WHO’s GPELF recommendations(6,7). Following effective implementation of MDA globally, at risk population has reduced by 58.6% compared to 2010 and 33 countries out of 72 stopped MDA in 2023(8,9). In South East Asia Region (SEARO), five out of eight endemic countries have achieved validation for eliminating LF as a public health problem as of 2025. Out of the1089 endemic districts in the Region, 73% have stopped MDA and are under post-MDA or post-validation surveillance.

In Nepal, 25 million people were at risk in 64 LF endemic districts as of 2020 (10,11), which has been reduced to 5.3 million in 2024 accounting for 78.8% reduction after several rounds of MDA (12).

WHO recommends evaluating transmission impact of IDA-MDA, through conducting an Epidemiological Monitoring Survey (EMS) nine months after completing two effective rounds (≥65% epidemiological coverage), followed by an IDA Impact Survey (IIS)(7,9). In Nepal, the first and second round of MDA with IDA was administered in six endemic districts namely Jhapa, Dhanusa, Mahottari, Sarlahi, Rauthat and Rasuwa in 2023 and 2024 as per eligibility criteria of WHO Guideline(7,12). Our study aimed to assess the reduction of LF infection prevalence below the transmission threshold in the 6 districts and examine the factors associated with Ag and MF prevalence.

## Methods

### Survey design and population

We conducted a cross-sectional EMS using a multi-stage sampling design among people aged more than 20 years, according to WHO M&E guidance on monitoring and epidemiological assessment of MDA(9) considering that age group more than 20 years and above carries the highest microfilariae burden and increased sensitivity to detect ongoing LF community transmission.

We selected six districts where two rounds of IDA MDA had been completed with an effective epidemiological coverage of ≥65% at district level.

### Sampling design and sample size

We divided the selected districts into evaluation Unit (EUs), each with a population of not more than 500,000, as per WHO guidelines (7,9,11). The EUs were defined based on risk factors, including low MDA coverage in the previous years, high number of resistant populations, high numbers of reported clinical cases, or proximity to low coverage areas. Considering the population size in the selected districts, we established one EU in Rasuwa, two EUs respectively in Dhanusa, Mahottari, Sarlahi, and Rauthat and 3 EUs in Jhapa district.

We selected two spot check sites in each EU based on factors such as having high number of clinical cases, vector abundance, and poor epidemiological coverage of IDA MDA. The sites selected in the 12 evaluation units and 6 districts are shown in Table 1.

A total of approximately 300 samples per site, or 600 samples per EU, were required, amounting to around 7200 samples across 12 EUs. This sample size was calculated to detect a critical threshold of <2% with a 5% chance of Type I error and ∼75% power when the true prevalence is 1%, accounting for a design effect of 1.5 for populations under 2,800 or 2.0 for populations over 2,800, in line with WHO M&E guidelines.

We calculated the required number of households to achieve a sample size of 300 adults (population above ≥20 years) per site, accounting for an expected 25% non-response rate, and an average of 2.65 adults per household(11). This resulted in the selection of approximately 151 households per site. We divided each site into equal segments containing approximately 100 households and two segment was randomly selected from each site using a lottery method.

A team of enumerators with the help of local health workers and female community health volunteers visited the households within the first selected segments and asked all available adults aged ≥ 20 years to participate in the survey. Then, remaining participants (300-number of participants enrolled from first segment) were asked from the households of next randomly selected segment to participate one by one.

### Data collection process

The survey team along with trained laboratory staff visited each household to collect the information about participation in the last MDA and uptake of IDA. They conducted the LF rapid antigen test using the Standard Q Filariasis Antigen Test (QFAT). All antigen positives were then followed up on same day or following day for night blood collection for microfilaria examination. (Table 1)

### Diagnostic Tests

We utilized the highly sensitive and specific Standard Q Filariasis Antigen Test (QFAT) for detecting LF antigen(13–15), followed by microfilariae testing through microscopy for those who tested antigen-positive, to assess any potential transmission signals.

We read the results in 20 minutes. Each QFAT was labeled with a unique ID so that tests could be linked with demographic information and other variables collected. The scoring of the positive test was performed based on the colour intensity of the test line. QFAT Test scores were recorded as negative if no test line was visible; and as positive if the test line was present. Tests with no control line were considered invalid results. In this situation, the test was repeated. All individuals who tested positive with QFAT were further tested for microfilariae through night blood slide collection.

### Microfilariae Testing

We conducted the Microfilariae testing using microscopy to detect microfilariae among all antigen positive individuals(16).

#### Sample collection

Night blood samples were collected between 10:00 PM and 2:00 AM to cover the peak appearance of microfilariae in the peripheral blood(17). Following a finger prick on the side pad, 120 µL of blood was drawn using a calibrated capillary tube and spread as three parallel lines (≈20 µL each) on a pair of glass slides. Two duplicate slides were prepared per sample to enhance diagnostic sensitivity(18–20).

#### Slide preparation and staining

The slides were air-dried overnight and then placed in distilled water for 5 minutes to dehemoglobinize the blood following standard procedures(21). After air drying for 1 hour, the slides were fixed in methanol for 5 minutes and stained with a 1:50 dilution of Giemsa stock for 50 minutes. The slides were air-dried again before being read under a microscope using 10X and 40X lenses to detect microfilaria.

#### Quality Control

All microfilariae-positive slides and negative slides were re-read by a second reader.

### Data analysis

We calculated the prevalence of antigen-positive and microfilariae-positive for each site. We divided the number of antigen-positive participants by total number of valid participants tested in the site. Similarly, we divided the number of microfilariae-positive participants by total number of valid participants. The individuals who tested either positive or negative in rapid diagnostic test using QFAT were taken as valid participants. Tests with no control line were considered invalid results and not included in the study as a denominator. We estimated the MDA treatment coverage, with 95% confidence intervals (CI), for each site using a one-sample non-parametric binomial test.

We assessed the differences in age distributions between individuals with antigen and microfilariae positive and negative tests using Wilcoxon rank-sum tests (Mann Whitney U) through IBM SPSS statistics 25. We analyzed the sample distribution by age groups (20–29, 30–39, 40–49, 50–59, 60–69, more mobile (working) versus less mobile population groups, occupations: (1) agriculture and housewife, and (2) all other occupations, cross-border residents, and non-cross-border residents. We compared the positivity rates of LF antigenemia and microfilaremia and the percentage of microfilariae test positive out of those antigen positives. We sequentially performed Chi-square (χ^2^) tests and Relative Risk (RR) analyses for each factor to assess associations with (1) MDA treatment coverage and (2) the prevalence of filarial antigen or adult filarial worms. Circulating filarial antigen serves as a marker of live, recently dead, or dying adult worms in the body(22,23). We sequentially applied Fisher’s Exact test and Relative Risk (RR) to evaluate the association of each factor with microfilariae (MF) prevalence.

We performed the multivariable logistic regression to calculate adjusted odds ratios (AORs) and examine the factors associated with the high antigen prevalence and microfilariae positivity.

### Ethical Consideration

The Ethical approval was not required, as this is the regular survey under National LF Elimination program and exempted by Nepal Health Research Council (NHRC) from the ethical review process (**Ref. no 1530 NHRC, December 10, 2020**). Participant confidentiality was ensured during the data collection. Written informed consent was obtained from all participants. Individual test results were communicated to the participants and public health action taken accordingly.

## Results

A total of 7,443 participants were recruited from 24 spot-check sites across 12 evaluation units (EUs) in six districts. 66% were female, and 25% were aged 20–29 years. The median age was 40 years (IQR: 29–55) and the mean age of participants was 43.24 years.

The LF antigen prevalence across the 24 surveyed sites in 12 evaluation units (EUs) was 4.47%, and microfilaria (MF) prevalence was 0.34%, with heterogeneity between sites. The antigen-to-MF ratio (Ag:Mf) was 13:1. Nine of 12 EUs: Jhapa A, Jhapa C, Dhanusa A, Dhanusa B, Mahottari A, Mahottari B, Sarlahi A, Rauthat A, and Rasuwa met the passing threshold (MF <1% at both spot-check sites). Three EUs (Jhapa B, Sarlahi B, and Rauthat B) failed, as at least one site in each exceeded the MF threshold.

Participants with antigen-positive had a higher mean age, exceeding antigen-negative participants by 3.2 years (46.3 vs. 43.1 years), p = 0.002. Similarly, microfilaria (MF)-positive individuals were older, with a mean age of 52.76 years, compared to 43.21 years among MF-negative participants (p = 0.003) (Table 2).

Three factors were found to be associated with antigen positivity. Female sex remained protective (AOR: 0.564, 95% CI: 0.441–0.721; *p*<0.001). Residing in border areas was associated with higher odds of antigen positivity (AOR: 1.754, 95% CI: 1.398–2.201; *p*<0.001). Receiving MDA in 2024 was protective (AOR: 0.554, 95% CI: 0.398–0.772; *p*<0.001). (Table 3).

Three factors were associated with MF positivity. Older age (≥40 years) was significantly associated with higher odds of MF positivity (AOR: 0.211 for 20–39 years vs. ≥40 years, 95% CI: 0.071–0.626; *p*=0.005). Male sex was a significant risk factor (AOR: 0.326 for females vs. males, 95% CI: 0.129–0.826; *p*=0.018). Receipt of MDA in 2024 has protective effect (AOR: 0.089, 95% CI: 0.017–0.464; *p*=0.004). (Table 4).

Four factors were associated with the MDA uptake: age, gender, cross-border residents and occupation. Participants aged <40 years were less likely to receive MDA treatment than those ≥40 years (AOR = 0.72, 95% CI: 0.653–0.793, p<0.001). Females had higher coverage than males (AOR = 1.438, 95% CI: 1.29–1.603, p<0.001). Cross-border residents were less likely to be treated than non-cross-border residents (AOR = 0.616, 95% CI: 0.558–0.681, p<0.001), whereas individuals in agriculture and housewife occupations had higher coverage compared to other occupations (AOR = 1.144, 95% CI: 1.008–1.298, p=0.037) (Table 5).

## Discussion

Our survey found the LF antigen prevalence at 4.47% and microfilaria prevalence at 0.34%, female sex and receipt of MDA in 2024 were protective against antigen positivity, while residence in border areas was associated with higher antigen prevalence. The microfilaria positivity was associated with older age (≥40 years), male sex, and lack of participation in recent MDA. The MDA uptake in 2024 varied significantly by age, sex, border residence, and occupation, with lower coverage among younger adults, males, and cross-border populations. These findings suggest that while active LF transmission is low and focal, residual infection persists, and gaps in recent MDA coverage, particularly in specific demographic and geographic groups, remain critical barriers to elimination.

### LF antigenemia and microfilaremia prevalence

The observed antigen prevalence of 4.47% alongside a much lower microfilaria prevalence of 0.34% indicates that adult *Wuchereria bancrofti* infections remain detectable, but active transmission is limited. This result is comparable with the EMS conducted in 5 districts/11 EUs in (24,25) early 2024 in Nepal (Ag prevalence 4.78% and Mf prevalence 0.35%). The Ag:Mf ratio of approximately 13:1 is consistent with previous studies conducted in post-MDA settings and reflects both parasite biology and treatment effects. Antigen positivity can persist for years even after rapid clearance of microfilariae following MDA, particularly due to presence of adult worm beyond its reproductive age. Antigen can also be detectable without any presence of Mf when infections involve single-sex or worms of either sex that do not produce microfilaria(25–29). This biological lag likely explains why recent MDA had a much stronger protective effect on microfilaremia than on antigenemia.

Despite antigenemia exceeding the 2% threshold in 16 of 24 sites, only three sites crossed the WHO microfilaria threshold of 1%, suggesting that ongoing transmission is limited despite the wider presence of adult worm. These findings support that absence or very low levels of microfilaremia in adult population is a strong indicator of interrupted transmission(30).

### Factors associated with antigen positivity

Female sex remained protective against antigen positivity, consistent with findings from other endemic setting(31,32). This may reflect differences in exposure patterns, health-seeking behavior, or higher MDA participation among women. Residence in border areas was associated with higher antigen prevalence, suggesting persistent exposure or delayed clearance of adult infections in these populations. However, this association likely reflects programmatic factors, particularly lower MDA coverage rather than intrinsic transmission risk alone. Receipt of MDA in 2024 was protective, reinforcing the effectiveness of recent treatment in reducing infection reservoirs, even if antigen clearance lags behind microfilarial clearance(33–36). For LF programs, these findings underscore the importance of sustaining high MDA coverage in hard-to-reach and mobile populations to reduce residual antigenemia over time.

### Factors associated with microfilaria positivity

Microfilaria positivity was associated with older age (≥40 years) and male sex, patterns widely reported in LF-endemic settings(37–40). Older age likely reflects cumulative exposure and longer duration of untreated infection, while higher prevalence among men may be related to occupational exposure or lower MDA compliance. Recent MDA participation emerged as the protective factor against microfilaremia, with a markedly lower adjusted odds compared to antigen positivity. This finding aligns with evidence from Samoa, India, and Kenya demonstrating rapid reductions in microfilariae following recent treatment rounds (41–43). These results emphasize that microfilaremia is highly sensitive to recent program performance and can serve as an early indicator of gaps in MDA delivery.

### LF transmission associated with Cross-border residence

Three evaluation units failed EMS due to sites exceeding the WHO microfilaria threshold, and most of these sites were located beside the international border. While unadjusted analyses suggested higher infection risk among cross-border residents, this association was attenuated after adjusting for recent MDA participation. This indicates that elevated infection in border areas is more likely driven by lower treatment coverage and cross-border movement during mass drug administration. Similar observations have been reported among migrant and mobile populations in Togo and Côte d’Ivoire, where infection risk is largely mediated by missed or incomplete MDA exposure(44–46).

### Determinants of MDA uptake

The analysis of 2024 MDA coverage revealed systematic disparities by age, sex, residence, and occupation. Younger adults (<40 years) were significantly less likely to receive treatment, consistent with studies from India and Pacific Island countries, where mobility, work-related absence, and lower perceived risk reduce compliance (41,45). Females had higher coverage than males, likely reflecting greater availability during household-based distribution and stronger health-seeking behavior, as documented in Nepal and Ghana (47,48). Cross-border residents had substantially lower coverage, reinforcing the need for targeted interventions in mobile populations(49). Higher uptake among individuals engaged in agriculture or household roles suggests that individuals more settled in their communities are more accessible during MDA campaigns(50,51). These patterns are directly relevant for program refinement, as persistent pockets of low coverage can sustain transmission even when overall indicators improve.

## Limitations

Our study has three major limitations: first the cross-sectional design limits causal inference, second self-reported MDA participation may be subject to recall bias, and third the antigen persistence limits interpretation of antigen prevalence as a marker of current transmission.

## Conclusion

This survey led to three conclusions. First, nine out of 12 evaluation units (EUs) had lymphatic filariasis (LF) infection measured as microfilaria (MF) prevalence, below the WHO threshold of 1% and successfully passed the EMS. Second, MF prevalence was strongly influenced by participation in the recent round of MDA. Third The MDA varied according to age, sex, cross-border residence, and occupation. Based on these conclusions, we propose the following recommendations. First, Nepal to conduct the IDA impact survey in the nine EUs that successfully passed the EMS—representing a further step in the national LF elimination program. Second, we should sustain effective MDA rounds in the evaluation units which exceeded the WHO threshold. Third, we should focus MDA rounds on male population, older adults, mobile young population, and cross-border residents. Fourth, additional targeted strategies are needed, such as synchronized cross-border MDA, tailored social mobilization, and flexible delivery approaches to ensure equitable treatment coverage in border communities. Overall, these recommendations will contribute to further reducing the LF transmission and achieving the elimination status.

## Contributors

RKM, RK conceived and designed the manuscript; RKM, GD, SRP, RK, and CBJ managed and supervised the data collection; AC and DS re-examined all the positive and Negative slide of MF to assure the quality. RKM, BB, SRP wrote the original draft., RN, CBJ, KB, PS, GD, BPD, BPD, BB, KPP, SK, reviewed and revised the manuscript. All authors read, reviewed, and approved the final manuscript.

## Declarations of interests

The authors declare no competing interests, and the views expressed in this report are solely those of the authors and do not necessarily reflect the positions of their respective organizations.

We extend our sincere gratitude to all Provincial Health Directors, Chiefs and LF focal persons of Health Offices, section chiefs, and municipal focal persons from the areas where the EMS was conducted for their valuable cooperation, coordination, and supervision. We also express our deep appreciation to the VBDRTC and EDCD teams for their significant contributions to the coordination and monitoring of the survey activities. We sincerely thank the data collectors and counselors for their dedication and commitment to quality data collection. Our heartfelt gratitude goes to Mr. Shivjee Das of the Health Office, Dhanusa, for his coordination, support, and onsite coaching during field data collection. Finally, we acknowledge the invaluable contributions of the trained microscopists: Mr. Aman Kumar Chaudhary (Kathariya PHC, Rautahat), Mr. Yub Nath Baral (Health Office, Jhapa), Mr. Roshan Sapkota (Gaurigunj PHC, Jhapa), Mr. Sanjay Kumar Sah (Gaushala Nagar Hospital, Mahottari), Mr. Bhogendra Kumar Yadav (Sabila Health Post, Dhanusa), Mr. Shambhu Prasad Kushwaha (Health Office, Dhanusa), Mr. Narendra Yadav (Mahottari Public Health Laboratory, Mahottari), Mr. Bhupendra Sahni (Achalgadh PHC, Sarlahi), Mr. Bishwajit Jha (Pataura Health Post, Rautahat), and Mr. Ram Saran Poudel (Thulogau Health Post.

## Data availability

The datasets generated and/or analyzed during the current survey are not publicly available to safeguard the privacy of individual patients. The data are available from the corresponding author upon reasonable request.

